# Artificial Intelligence Powered Research Automation (AIPRA) Versus Human Expert: A Two-Arm Ophthalmology Comparative Study

**DOI:** 10.1101/2025.10.27.25338904

**Authors:** Ayman Musleh, Nouran Alwisi, Hashem Abu Serhan, Ahmad Toubasi, Lna Malkawi, Saif Aldeen Alryalat

## Abstract

**Purpose:** To compare the quality and efficiency of an AI-powered research automation (AIPRA) workflow with a conventional human-led workflow for producing a full systematic review manuscript on the same question.

**Methods:** Two independent pipelines (human-led vs. AIPRA) each generated a complete manuscript addressing “What is the role of large language models in glaucoma diagnosis?”. No protocols or templates were shared. Three blinded domain experts rated five domains on 5-point Likert scales. The primary endpoint was the overall quality of each workflow from query to final manuscript.

**Results:** Mean total scores: human 74.7%, AIPRA 65.3%. The mean difference (AIPRA - Human) was −9.3% (95% CI, −18.8% to 0.0%), meeting the pre-specified non-inferiority criterion. Domain means were identical for query development (66.7% each); the human-led pipeline scored higher in screening, field selection, full-text extraction, and manuscript writing. AIPRA completed the workflow in approximately 2 hours versus about 1 month for the human pipeline (375x faster).

**Conclusion:** AIPRA was non-inferior to human experts on overall quality while drastically reducing time to completion. Appropriate human oversight remains important, especially for screening and extraction tasks.

## 1. Introduction

Systematic reviews are a fundamental component of evidence-based medicine. They provide clear and reproducible syntheses that help with clinical decisions, guideline creation, and health policy development (1). In the rapidly advancing field of ophthalmology, systematic reviews are important for combining the growing amount of diagnostic, imaging, and treatment information. By bringing together different findings, these reviews reduce research overlap, help with evidence-based decision-making, and clearly point out ongoing gaps in knowledge. Conventional systematic review workflows are time-consuming and can be labor-intensive, often struggling to keep pace with the rapidly evolving literature (2). They often require weeks or months for manual title and abstract screening, retrieving full texts, extracting data, assessing bias, and synthesizing narratives. Variations among reviewers and the possibility of inadvertently overlooking relevant studies further complicate this process. In a study, the cost of associated labor in a systematic review is approximately $140,000 where automation (e.g., machine learning, artificial intelligence) could significantly lower the cost (3).

The introduction of artificial intelligence (AI) technologies, particularly large language models (LLMs), has created new possibilities for automating many of these tasks. LLMs can now perform literature searches, extract data from full-text papers, comprehend complex clinical questions, and write scientific manuscripts, in addition to demonstrating language proficiency (4,5). Initial reports suggest that LLM-assisted workflows may enhance efficiency (6–10). However, there is still a lack of rigorous, head-to-head comparison between human-led and AI-driven systematic reviews across the full methodological spectrum. Rapid advancements in imaging, biomarkers, pharmacology, and AI-driven diagnostics have revolutionized the research and management of glaucoma. As a result, synthesizing evidence in a timely and accurate manner has become more challenging.

This study examines the role of AI and LLMs in enhancing systematic review workflow quality and efficiency by comparing an AI-powered research automation (AIPRA) systematic review with a conventional human-led systematic review approach. The literature search, study selection, data extraction, synthesis, and manuscript generation were all carried out independently by each method. The comparison in the quality of both manuscripts was done by blinded experts using a predefined criteria. We hypothesized that integrating AI into evidence synthesis would be non-inferior to human reviewers in overall quality while markedly improving time efficiency.

## 2. Materials and Methods

### 2.1. Study design

We conducted a two-arm, parallel comparative study in which independent pipelines: (i) a human-led review team and (ii) an AI-powered research automation (AIPRA) workflow were compared. Each generated a full systematic review manuscript addressing the same research question: “What is the role of large language models in glaucoma diagnosis?”. No shared protocol, eligibility template, search strategy or reporting framework was provided. Both pipelines were given only the research question. Each manuscript’s Methods section served as its self-documented record of search, selection, extraction, and appraisal procedures. No reconciliation or harmonization between pipelines was attempted. The resulting manuscripts were subsequently masked and evaluated by three expert raters. Both Manuscripts are provided in the supplementary material.

### 2.2. Comparison arms

#### 2.2.1 Manuscript Generation

The AIPRA workflow was performed using the “High Yield Med” platform (https://platform.highyieldmed.org/). The platform is an end-to-end solution that provides the user to conduct the systematic review within it. It has an implemented step-wise automation, combined with LLM generation and human in the loop at each step to confirm the output before moving to the next step. The workflow in each step is as follows: (1) Search Query: The platform utilizes an internal model to dissect the research question into different topics, that are then used to build a search query for each with an LLM agent. Another agent then validates the query for the topic by assessing its search results and modifying accordingly. The query developed from each group of agents are then combined into the final search query for the selected database (i.e., PubMed in this project). (2) Title/abstract screening: The platform retrieves the articles’ metadata based on the query and feed them an LLM agent that screen them based on the inclusion and exclusion criteria, providing the decision to include or exclude and the reason for the decision. This is followed by a human confirmation before moving to the next step. (3) Fields to extract: Based on the research question and sample included articles, another agent constructs the fields to extract from full text studies. (4) Extraction table: the platform constructs the full text extraction table, where each of the extraction fields will be filled by an agent for each study. At this point, another screening process occurs to exclude articles not meeting the inclusion criteria. This is different than the human led projects, where this step happens before the extraction table. (5) Full text drafting: This is a multiagent process, where there are five different agents working to draft the manuscripts based on the extraction table and the research question. These agents are equipped with tool calling to retrieve relevant articles when drafting the introduction and discussion.

While the platform automates the processes and integrates AI within each step, expert human interaction is required to confirm and proceed to the next step. For the current project, we limited human experts to confirming study eligibility and data extracted, acting as “The second reviewer” in the process, and the interventions were documented in detail. Less than 3% of the extracted content was edited by the human expert to complement the data extraction, which were 9 fields from the total of 306 fields within the extraction table. Otherwise, no human intervention allowed in the query or drafting.

#### 2.2.2. Human-led review

The human-led manuscript was produced according to the expert’s routine evidence-synthesis practices. This was led by two authors (AM & NA) that conducted the query generation, screening articles, decided on field extraction, performed the data extraction, and drafted the manuscript. No LLM used in the process except for grammatical corrections and rephrasing in manuscript writing.

### 2.3. Evaluation and Rating

The evaluation tool included five domains that represent the pertinent and expert needed steps in systematic review, including query development, screening quality, field selection for data extraction, full-text data extraction, and manuscript structure. Each domain included a 5-point Likert scale (1 = poor, 5 = excellent). **Table 1** details the evaluation criteria used to assess the quality of each pertinent step in the systematic review done. In addition to quantitative scoring, raters provided qualitative feedback in the form of free-text global comments on strengths and weaknesses.

**Table 1:** Evaluation criteria used to assess the quality of each pertinent step in the systematic review done.

|  | **Query Development** | **Screening of Searched Articles** | **Field Selection for Data Extraction** | **Full Text Data Extraction** | **Manuscript Writing** |
| --- | --- | --- | --- | --- | --- |
| **Score 1 (Poor)** | Search query lacks relevant terms, operators, or truncations; unrelated articles dominate results; query is either too broad or too narrow without justification. | Inconsistent inclusion/exclusion; reasons missing or irrelevant; skipped articles without justification. | Chosen fields do not align with study aim; missing key variables. | Unable to extract relevant data. | Poor structure; major missing sections; inaccurate or hallucinated content; ignores PRISMA guidelines. |
| **Score 2 (Fair)** | Includes some relevant operators and terms but limited optimization; sensitivity or specificity is noticeably imbalanced. | Most reasons are relevant but inconsistent; some articles not screened. | Some fields relevant but missing important ones. | Extracts some but not all key data. | Some structure present but with errors; limited data integration; partial PRISMA adherence. |
| **Score 3**<br>**(Good)** | Uses PubMed operators/truncations adequately; query retrieves a reasonable mix of relevant studies with acceptable sensitivity/specificity. | Clear, mostly correct reasons for inclusion/exclusion; nearly all articles screened. | Most selected fields are relevant and aligned with aim. | Extracts main data correctly. | Well-structured manuscript with most sections complete; mostly correct data; good PRISMA adherence. |
| **Score 4 (Very Good)** | Well-structured query with most relevant terms; strong use of Boolean operators, filters, and truncations; balanced sensitivity and specificity. | Consistently applies correct inclusion/exclusion criteria; all articles screened; reasons are clear. | Fields fully reflect aim with minor omissions. | Data extraction is accurate with minimal errors. | Strong structure, accurate use of prior data, minimal hallucinations; follows PRISMA closely. |
| **Score 5**<br>**(Excellent)** | Fully optimized query with precise operators, MeSH terms, and filters; highly relevant results with | Perfect consistency and accuracy in inclusion/exclusion decisions; detailed and correct reasons for every | All fields are directly aligned with aim; no omissions; clear rationale for each. | Comprehensive and accurate extraction from all articles. | Fully polished, well-structured manuscript; all data accurately used; zero hallucinations; PRISMA perfectly followed. |
|  | minimal<br>irrelevant<br>articles. | article. |  |  |  |

We recruited three independent domain experts with backgrounds in systematic reviews and meta-analyses, who were blinded on the workflow used for each project as well as to each other’s evaluations. Each reviewer was given each manuscript, the data extraction table generated by each workflow, and the full text articles included for both workflows.

### 2.4. Primary and Secondary Outcomes

The primary endpoint was the overall quality of each workflow from query to final manuscript, which was calculated as a percentage (combined scores divided by 25). We defined a non-inferiority margin of 80% quality of human-led manuscript, meaning that the AIPRA workflow quality should not fall more than 20% of what human-led workflow had (11). Secondary endpoints included time required to complete each step for each workflow, and the amount of human intervention needed for the AIPRA workflow. We also performed a qualitative assessment through the reviewer’s interview on each project’s quality.

### 2.5. Statistical Analysis

A paired non-inferiority analysis was performed to compare the overall quality scores between the AIPRA and human-led systematic review workflows across five evaluation domains. For each domain, the difference in percentage score (AIPRA–Human) was calculated, and the mean difference along with its 95% confidence interval (CI) was determined. The standard error of the mean difference was estimated as the sample standard deviation divided by the square root of the number of paired observations. A two-sided 95% CI was constructed using the Student’s *t*-distribution. A non-inferiority margin of −20% was pre-specified, meaning that AIPRA would be considered non-inferior if the lower limit of the 95% CI for the mean difference did not cross −20%. Statistical computations were conducted using standard methods in Python (SciPy package).

## 3. Results

### 3.1. Overall Manuscript Quality

Across all raters, the human-led manuscript achieved a mean total score of 74.7% (range 64–84%), whereas the AIPRA-generated manuscript received a mean score of 65.3% (range 36–96%). Across the five evaluated domains, the AIPRA workflow demonstrated a mean quality difference of −9.3% compared to the human-led workflow (AIPRA–Human), with a 95% confidence interval of −18.8% to 0.0%. Although AIPRA scored slightly lower on average, the lower bound of the confidence interval remained above the predefined non-inferiority threshold of −20%, indicating statistical non-inferiority. In other words, the AIPRA-generated systematic review preserved more than 80% of the overall quality achieved by human reviewers while substantially reducing completion time. **Figure 1** shows the domain level score for both workflows.

**Figure 1.**
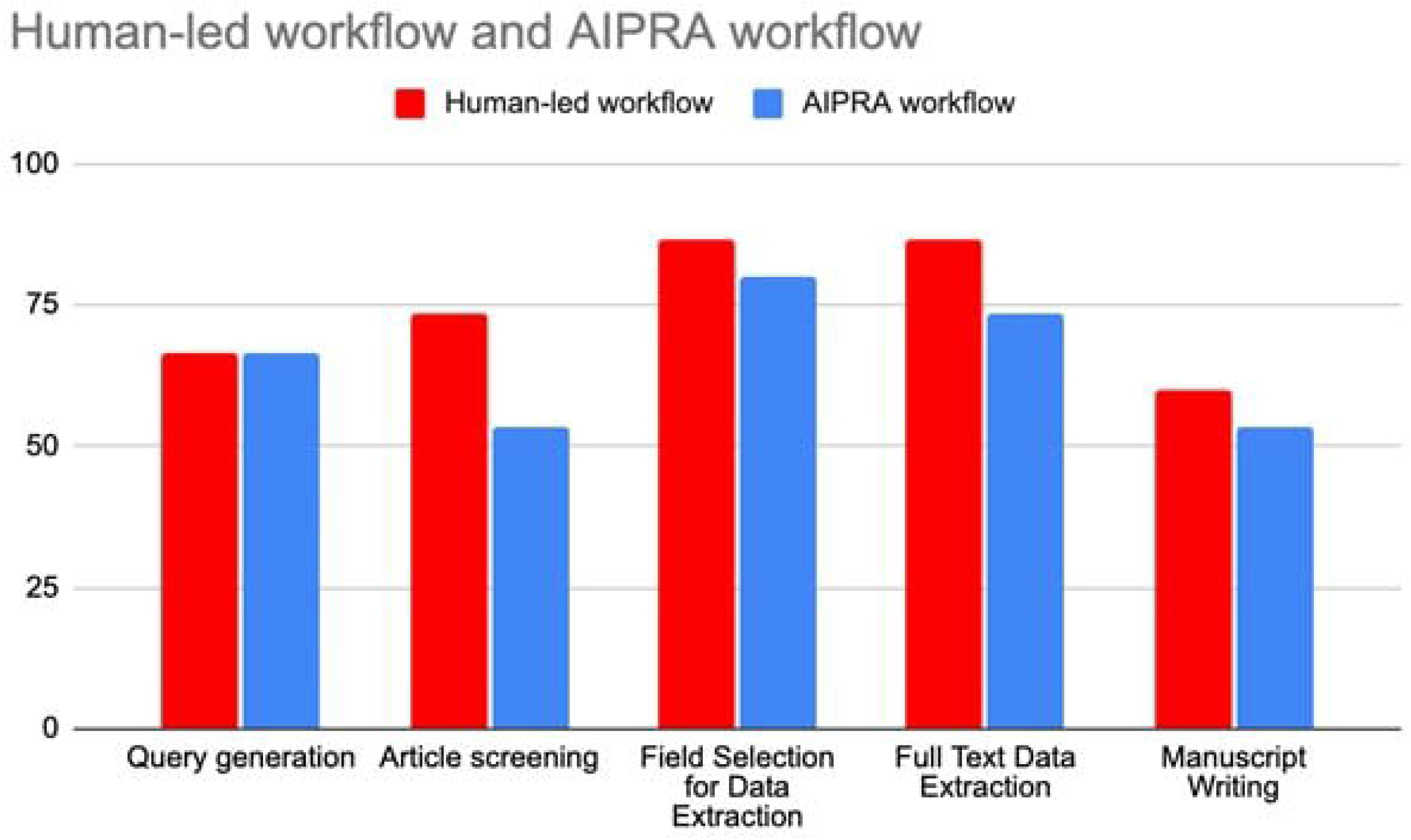
Domain-Level Comparison of Quality Scores Between AIPRA and Human-Led Workflows.

Mean scores across the three raters were identical for query development (66.7% for both). The human-led manuscript achieved slightly higher scores in screening quality (73.3% vs. 53.3%), field selection for data extraction (86.67% vs. 80%), full-text data extraction (86.67% vs. 73.33%) as well as manuscript structure (60% vs. 53.33%).

### 3.2. Time Efficiency

A substantial difference was observed in the total time required to complete each workflow. The AIPRA workflow produced a complete review in approximately two hours. In contrast, the human-led team required approximately one month to complete the same process manually using conventional tools. This reflects an estimated 375X less time, highlighting AIPRA’s potential to drastically compress review cycles.

### 3.3. Qualitative Appraisal

The human-led manuscript was praised for its clear structure, logical flow and strong adherence to PRISMA guidelines. Reviewers highlighted the thoroughness of field selection and data extraction, although one noted that the inclusion of detailed performance metrics extended beyond what was necessary for the study’s primary objectives. Suggested areas for improvement included expanding the background section, clarifying the study aims as well as enhancing methodological detail. The AIPRA-generated manuscript was commended for its well-constructed search strategy, which retrieved a broad and relevant set of studies, and for the strong alignment between selected variables and the research objectives. However, reviewers also identified limitations, including inconsistent use of abbreviations, non-inclusion of some relevant studies, and sections where the narrative structure and language could be more formally scientific.

## 4. Discussion

In this two-arm comparative study, the AIPRA workflow was non-inferior to human led workflow, completing a full systematic review with all its pertinent steps with 375X less time and effort needed through a human led systematic review. The AIPRA workflow integrates LLMs in a controlled and structured manner, utilizing them only for predefined, labor-intensive tasks that can be safely automated. Through a multi-agent architecture, built-in quality assurance measures, and human-in-the-loop oversight, AIPRA accelerates the systematic review process while maintaining the reliability and scientific rigor of the final output.

Since the introduction of large language models (LLMs) such as ChatGPT in late 2022, their adoption in scientific research has been both rapid and inevitable (12,13). Initially, their application was primarily focused on manuscript drafting and editing, given their strong performance in language-related tasks. However, misuse by some researchers (14). prompted concerns about transparency and integrity in scientific writing. In response, multiple organizations have issued and continuously updated guidelines to regulate and standardize the responsible use of LLMs in research and publication practices (15,16). The AIPRA framework was designed to leverage the advantages of LLMs while maintaining strict adherence to emerging ethical and scientific guidelines for their use. Each component of the workflow incorporates predefined control points that restrict LLM activity to specific, auditable tasks.

The pattern found in our analysis aligns with emerging literature showing that LLMs achieve high specificity and competitive accuracy in screening while being markedly faster and cheaper than manual review. (8, 17–18) Oami et al prospectively evaluated GPT-4 Turbo for title/abstract screening in five clinical questions from the Japanese sepsis and septic shock guidelines. (17) Using human full-text included studies as the reference, the LLM achieved pooled sensitivity of 0.75 and specificity of 0.99. Also, the LLM was ∼13× faster than humans, taking 1.30 minutes per 100 records versus 17.2 minutes for humans. (17) Li et al. systematically benchmarked multiple large language models (ChatGPT-4/3.5 and later 4-Turbo, Gemini 1.0 Pro, Llama 2/3, Claude 3 Opus, and PaLM 2) for abstract screening across three public “SYNERGY” datasets. (8) Top-performing models achieved balanced sensitivity and specificity, with overall accuracies approaching or exceeding 0.90, though performance varied by dataset and model version. The authors also reported notable efficiency and affordability; for example, screening 200 abstracts cost approximately $6 for GPT-4.0, $0.2 for GPT-3.5, and $10 for Llama 2. (8) Processing the same dataset typically took 10–20 minutes on a single thread. Overall, both the monetary and time costs of automated abstract screening were minimal compared with manual review, making LLM-based workflows a highly scalable and cost-effective alternative. Beyond screening, recent studies suggest LLMs can reliably perform data extraction. One evaluation reported 92.4% concordance with human consensus and 94.1% reproducibility between independent AI sessions, with most discrepancies occurring when source texts lacked explicit data. (18)

A major strength of this study lies in its controlled, masked, head-to-head design directly comparing human experts and an AI system in the generation of systematic reviews, both addressing an identical research question with a clearly defined primary endpoint. This design enabled a transparent evaluation of automation performance. Nonetheless, several limitations should be considered when interpreting our findings. First, both reviews relied on a single database. Furthermore, unlike standard systematic review which routinely include quality assessment and risk of bias appraisal; neither the human-generated nor the AI-generated reviews in this study conducted such evaluations.

Future research should incorporate larger and more heterogeneous panels of blinded raters to enhance scoring precision and improve the generalizability of findings across different domains. Also, establishing standardized reporting frameworks for AI-assisted systematic reviews would clarify the boundaries of automation and facilitate transparent peer evaluation.

In conclusion, our findings highlight the transformative potential of AI-assisted evidence synthesis. Through the AIPRA workflow, the systematic review showed non-inferiority to expert human-led systematic review with a fraction of time and effort, without compromising the integrity of the research. When appropriately guided by expert oversight, AIPRA systems can drastically accelerate systematic review production without substantial compromise in accuracy or scholarly quality. Continued advancements in AI model governance, transparency, and reporting standards will further strengthen their role in future evidence-based medicine.

## Supporting information

Human-led Data extraction Table

Human-led Manuscript

AIRPA Data extraction Table

AIRPA Manuscript

## Data Availability

All data produced in the present study are available upon reasonable request to the authors

## Acknowledgement

This article was partly revised by ChatGPT-4o, a large language model from OpenAI. The model was only used to improve the readability and language quality of the article. The version of ChatGPT-4o used was gpt-40-2025-10-18. It was used in several parts of the article, including but not limited to the introduction and discussion. It was used under strict human supervision and control. Additionally, authors carefully reviewed and polished the content generated by the model. The model was only used to improve the language and readability of the article and was not used to copy or replace any researcher tasks, such as generating scientific insights, analyzing and interpreting data, or drawing scientific conclusions.

