## Supplementary material for "Artificial Intelligence Powered Research Automation (AIPRA) Versus Human Expert: A Two-Arm Ophthalmology Comparative Study": Human-led Manuscript

Manuscript 1

1. **Introduction**

Glaucoma is a group of progressive optic neuropathies characterized by irreversible damage to the optic nerve and corresponding visual field (VF) loss. (1) It is a leading cause of irreversible blindness worldwide, often progressing insidiously without noticeable symptoms until advanced stages.(1) Because glaucoma damage is permanent, early detection and intervention are critical to prevent vision loss. Artificial intelligence (AI), particularly machine learning and deep learning with convolutional neural networks (CNNs) have shown promise in disease diagnosis.(2,3) Numerous algorithms have been trained on retinal fundus photographs and OCT scans to automatically detect changes in the optic nerve. These AI systems can analyze features like the optic cup-to-disc ratio, nerve fiber layer thickness, and VF patterns at a speed and consistency beyond human capability. (4)

Large Language Models (LLMs) represent a different paradigm in AI for healthcare. They are a class of AI models trained on enormous texts to understand and generate human-like language. In contrast to CNNs that excel at image analysis, LLMs specialize in natural language processing and they can interpret text input, answer questions, summarize information, and even engage in conversation using medically relevant knowledge.(5)The distinction between image oriented AI and language models is beginning to blur with the development of multimodal systems. New frameworks are being developed to combine vision and language AI, enabling analysis of ophthalmic images alongside textual data In principle, a multimodal LLM-based system could take in a fundus photograph, an OCT report, and a patient’s history, and then generate an integrated diagnostic assessment or recommendation. (6)

In this review, we aimed to systemically evaluate the role of LLMs in glaucoma diagnosis.

2. **Methods**

**2.1. Protocol**

We performed this meta-analysis in accordance with the Preferred Reporting Items for Systematic Reviews and Meta-Analysis (PRISMA).

**2.2. Literature search**

We conducted a systematic search on the PubMed database on August 24, 2025, to identify relevant studies. The following search strategy was applied: ("Large Language Models"[Mesh] OR "Generative Artificial Intelligence"[Mesh] OR "large language models" OR LLM OR LLMs OR ChatGPT OR "Chat-GPT" OR "Chat GPT" OR GPT OR LLaMA OR Bard OR Gemini OR deepseek) AND ("Glaucoma"[Mesh] OR "Ocular Hypertension"[Mesh] OR Glaucoma OR "Ocular Hypertension").

**2.3. Eligibility criteria**

We included studies that: 1) evaluated the use of LLMs in glaucoma diagnosis, 2) were original primary research articles (excluding reviews, conference abstracts or editorials) and (3) were published in English. Studies that did not meet all these criteria were excluded.

**2.4. Study selection**

The articles retrieved from the search were screened using title/abstract then the remaining studies were screened using their full-text form.

**2.5. Data collection process**

The following variables were collected: study title, year of publication, study design, LLM used, dataset, reference standard, sample size, and diagnostic performance metrics (sensitivity, specificity, accuracy, and AUC). The complete data extraction table is provided in the **supplementary materials.**

3. **Results**

**3.1.Overview**

Our database search identified a total of 117 records, all retrieved from PubMed. After title and abstract screening, 93 records were excluded, leaving 24 articles for full-text review. Following detailed eligibility assessment, 11 reports were excluded (6 due to wrong outcome and 5 due to inappropriate study design). Ultimately, 13 studies met the inclusion criteria and were incorporated into the systematic review. **Figure 1** provides a detailed summary of the search and selection process.

**Fig. 1**. Study selection flow chart, Preferred Reporting Items for a Systematic Review and Meta-analysis

**3.2 Study characteristics**

Thirteen studies published between 2023 and 2025 were included in this systematic review, encompassing research from seven countries. The United States contributed the largest number (n=6), followed by China (n=2), and single studies from Singapore, Jordan, the United Kingdom, Japan, and Australia. Study designs varied in data modality: most relied on textual case reports (n=7), while others focused on image-based inputs using color fundus photographs (CFPs) (n=5) or VF reports (n=1). One study was exclusively VF based. Across the 22 LLM models reported, the most frequently evaluated were ChatGPT-4.0 (n=6), ChatGPT-4V (n=4), ChatGPT-3.5 (n=3) and ChatGPT-4o (n=3). Single-study appearances included GPT-4.5-preview, DeepSeek-R1, DeepSeek, ChatGPT-01, Qwen-2.5 MAX, and Google Gemini. Also seven of the included studies directly compared the diagnostic performance of LLMs with ophthalmologists.

**3.3. LLMs Applied to Textual Case Reports**

In a large OHTS-based evaluation of 3,170 eyes from 1,585 participants, data from both eyes were converted into textual case reports and fed to ChatGPT for glaucoma diagnosis; GPT-4.0 achieved Area Under Curve (AUC) of 0.76, accuracy 87%, specificity 90%, sensitivity 61%, and F1 0.92, outperforming GPT-3.5. (7) Overall, GPT-4.0 provided higher diagnostic accuracy, whereas GPT-3.5 was more sensitive. Zhang et al. prospectively compared GPT-4o with three ophthalmologists on 26 glaucoma cases using text-only case summaries.(8) GPT-4o scored lower for the primary diagnosis. Aminan et al. introduced GlaucoRAG a glaucoma specialized retrieval augmented LLM. It wraps a GPT-4.5-preview with a glaucoma-specific knowledge base so answers are grounded in literature, guidelines, and textbooks. Using textual inputs only, it achieved 81.8% diagnostic accuracy on 11 case reports, vs 72.7% GPT-4.5 and 63.6% DeepSeek-R1; mean of three glaucoma specialists of around 66.7%. Ming et al. evaluated GPT-3.5/4.0 on 104 Chinese SRT ophthalmic cases. (9) They tested GPT models in the history-plus-examination (Hx+Ex) setting, and GPT-4.0 correctly identified all glaucoma cases placing glaucoma among the highest-performing subspecialties in the study.

**3.4. LLMs for Glaucoma Image Analysis**

Jalili et al evaluated GPT-4V on 300 disc-centered CFPs from ACRIMA, ORIGA, and RIM-ONE v3 with 139 glaucoma images and 161 non-glaucoma images. (10) They were analyzed using a standardized prompt via API and compared with two expert graders. GPT-4V achieved accuracy of 0.68%, 0.70% and 0.81% on (ACRIMA, ORIGA and RIM-ONE datasets and generally lower than experts. Srinivasan et al. evaluated GPT-4V and Gemini on 44 fundus photographs from the SEED study which contain 10 normal and 34 abnormal across six diseases including five glaucoma cases. (11) On the glaucoma subset, GPT-4V-default flagged 4/5 images as abnormal (80%), GPT-4V-Data-Analyst flagged 5/5 (100%), and Gemini flagged 1/5 (20%). However, all models performed poorly at disease-specific classification despite being able to flag abnormality. Using 12 fundus photos as image-only inputs with a standard prompt, Gupta & Al-Kazwini evaluated GPT-4.0 model. It correctly diagnosed 4/12 (33%), was partially correct in 1/12, and incorrect in 7/12. (12)The single glaucoma case was correctly identified, but errors and hallucinations were frequent. AlRyalat et al. evaluated ChatGPT-4 for glaucoma detection using the REFUGE test set. (13) The model, applied without fine-tuning, performed binary classification with an overall accuracy of 90.0%. Specificity was high at 94.4%, whereas sensitivity was markedly lower at 50.0% . Exploratory preprocessing of 200 images demonstrated a trade-off between sensitivity and specificity: cropping alone increased sensitivity to 87.5% but reduced specificity to 56.5%, while cropping combined with contrast limited adaptive histogram equalization achieved sensitivity of 62.5% and specificity of 55.4%. Tomita et al. tested GPT-4 models on 580 ophthalmic cases, finding higher accuracy with multimodal input: GPT-4o image+text 77.1%, GPT-4V image+text 71.0%, vs GPT-4V text-only 66.7%.(14) For glaucoma-specific items, accuracy reached 79.2%, with inputs combining clinical text plus images.

**3.5. LLMs Applied to Visual Field Data**

Tan et al evaluated GPT 4o on 60 Humphrey 24-2 VF reports and prompted to provide an overall VF-only diagnosis. (15)Across the full cohort, the model achieved 96.7% accuracy for the overall diagnosis from VF reports only.

**3.6. Comparisons Between LLMs and Ophthalmologists**

Several studies directly compared the diagnostic performance of LLMs with that of ophthalmologists and residents. Zhang et al. compared GPT-4o with three ophthalmologists for diagnostic performance. (8) GPT-4o was less accurate and less complete than all doctors for primary diagnoses. For differential diagnoses, however, GPT-4o matched Doctors A and C in accuracy, outperformed Doctor B. Overall, GPT-4o lagged behind clinicians in primary diagnosis but performed competitively in differential diagnosis, particularly in completeness. Huang et al., tested GPT-4 on 10 de-identified glaucoma cases rated by masked participants. The model’s case-based diagnostic accuracy was similar to that of fellowship-trained glaucoma specialists, with no statistically significant differences. (16) In contrast, Jalili et al. assessed GPT-4V on 300 disc-centered fundus photographs. The model demonstrated variable performance, occasionally achieving higher sensitivity but consistently showing lower specificity than expert graders.(10). Another study compared LLMs with three fellowship-trained glaucoma specialists and found that the models performed on par with the clinicians on the same case-report task. (17) GlaucoRAG achieved 81.8% (9/11), clearly outperforming specialists, GPT-4.5 reached 72.7% (8/11), modestly higher than specialists, while DeepSeek-R1 was 63.6% (7/11), slightly below specialist performance. Ming et al. evaluated GPT-3.5/4.0 vs residents on 104 Chinese SRT cases; in glaucoma, GPT-4.0 achieved 100% top-1 diagnostic accuracy with history+exam, while history-only triage to glaucoma was modest (60% vs residents 20%).(9) Likewise, Hussain et al. evaluated 11 glaucoma case reports from the University of Iowa. The diagnostic accuracy was 63.6% for both ChatGPT-4.0 and ChatGPT-01, 54.5% for DeepSeek-V3 and Qwen-2.5 Max, while human experts averaged 51.5% across three attending physicians). (18) Delsoz et al. evaluated ChatGPT-3.5 (text-only) and found 72.7% provisional diagnostic accuracy, comparable to senior ophthalmology residents. (19) See **Table 1**.

4. **Discussion**

LLMs can achieve promising but variable diagnostic performance for glaucoma, with reliability highly dependent on the input modality and use-case. Overall, our results suggest that LLMs perform best when provided with rich textual or multimodal clinical data. Notably, one study showed exceptional performance on VF reports as GPT-4.0 correctly classified 96.7% of 24-2 VF printouts as normal, suspect or glaucoma. (15). A recurrent theme is the trade-off between sensitivity and specificity. Higher-performing LLM versions tend toward higher specificity at the expense of sensitivity, whereas simpler models or adjusted inputs can raise sensitivity with more false positives. For instance, ChatGPT-4.0 was overall more accurate and specific than 3.5, but GPT-3.5 detected a greater proportion of true glaucoma cases with higher sensitivity of around 85% vs 61% albeit with many false positives. (7)

Similarly, GPT-4 Vision often flagged abnormal optic discs with high sensitivity but had lower specificity than expert ophthalmologists in image-based tasks.(10) Image preprocessing might shift this balance. For example, cropping fundus images around the optic disc dramatically increased GPT-4’s sensitivity from 50% to 87.5% at the cost of decreasing specificity.(13) Our review also reveals meaningful performance differences across LLM models and versions.

Generally,

OpenAI’s GPT-4 series demonstrated superior glaucoma diagnostic performance relative to other LLMs tested. For instance, GPT-4.0 consistently outperformed GPT-3.5 on the same tasks, (7). Non-OpenAI models have trailed in the limited evaluations so far. In a study of 11 complex cases, open-source models like DeepSeek and Qwen-2.5 MAX achieved only about 54–64% accuracy, substantially below GPT-4.0’s 63.6% and the reasoning ChatGPT-o1 model’s 63.6%. (18) Google’s multimodal Gemini model, tested on fundus photos, flagged far fewer glaucoma cases with only 20% sensitivity in one small series compared to GPT-4V which identified most abnormal discs. (11) These findings suggest that GPT-4 and its variants currently set the benchmark, while other LLM families remain less proficient in this domain unless further specialized or fine-tuned. Several studies showed that multimodal inputs and knowledge augmentation can markedly improve LLM diagnostic reasoning. (14)

Similarly, retrieval-augmented approaches have shown promise; Aminan et al. introduced GlaucoRAG, which couples GPT-4.5 with a glaucoma-specific literature database. Grounding the model’s answers in relevant guidelines and journal articles boosted diagnostic accuracy to 81.8%, outperforming the base GPT-4.5 model and even exceeding the average accuracy of glaucoma specialists on those cases. The authors noted that such knowledge grounding reduces hallucinations and improves clinical reasoning, albeit with added complexity.

LLMs have shown surprisingly strong performance in interpreting standard automated perimetry outputs, although evidence is limited. In one study using Humphrey 24-2 VF printouts, GPT-4 achieved 96.7% accuracy in categorizing eyes as glaucomatous, suspect, or normal based on the VF pattern alone. (15).

Our review covered a range of data modalities allowing us to integrate findings across diverse applications of LLMs in glaucoma diagnosis. We also gave special attention to studies that directly compared LLM performance with human clinicians which adds clinical context to the diagnostic metrics. Nonetheless, there are important limitations to acknowledge. First, our literature search was restricted to a single database, PubMed. It is possible that relevant studies indexed in other databases were missed. Second, the number of eligible studies was relatively small, and their heterogeneity was high. For example, OpenAI recently released GPT-5 with multiple modes, yet no published work to date has examined its role or potential impact in glaucoma diagnosis. We emphasize that our conclusions should be viewed as a current snapshot, with the expectation that more robust data will emerge to refine the understanding of LLMs in glaucoma care.

In conclusion, LLMs represent a novel and promising tool in glaucoma diagnosis. Current evidence imply that their capabilities vary widely by modality and context and at present their most appropriate role is as assistive tools for flagging abnormalities, informing decisions, and educating under close clinician oversight, rather than as autonomous diagnosticians. With ongoing improvements and careful implementation, LLMs could eventually become a valuable adjunct in glaucoma care.

**Table 1:**

| **Table 1** Performance comparison of LLMs and ophthalmologists in glaucoma diagnosis | | | | | |
| --- | --- | --- | --- | --- | --- |
| **Study** | **Models** | **Scale of accuracy** | **Results of the model** | **Results of the ophthalmologist(s)** | ***P* value** |
| Zhang, 2024 | ChatGPT-4o | 10-point Likert | 5.500 | Doctor A: 7.577 | <0.0001 |
|  |  |  |  | Doctor B: 6.808 |  |
|  |  |  |  | Doctor C: 8.038 |  |
| Huang, 2024 | ChatGPT-4 | 10-point Likert (reported as *mean rank* via Mann–Whitney U) | Glaucoma case accuracy: 197.4 | Glaucoma case accuracy of the specialist: 190.1 | 0.67 |
| Jalili, 2025 | ChatGPT-4V | The standard classification accuracy | ACRIMA Dataset: 0.68 | ACRIMA Dataset: Expert 1/Expert 2: 0.78/0.72 | <0.001 |
|  |  |  | ORIGA Dataset: 0.70 | ORIGA Dataset: Expert 1/Expert 2: 0.80/0.78 |  |
|  |  |  | RIM-ONE Dataset: 0.81 | RIM-ONE Dataset: Expert 1/Expert 2: 0.88/0.87 |  |
| Aminan, 2025 | DeepSeek R1 | The standard classification accuracy | 63.6% | Glaucoma Attending 1: 72.7% | NA |
|  | ChatGPT-4.5-PREVIEW |  | 72.7% | Glaucoma Attending 2: 63.6% |  |
|  | GlaucoRAG |  | 81.8% | Glaucoma Attending 3: 63.6% |  |
| Ming, 2024 | ChatGPT‑3.5 (hx only) | The standard classification accuracy | 1/5 (20%) | Residents: 1/5 (20%) | NA |
|  | ChatGPT‑4.0 (hx only) |  | 3/5 (60%) |  |  |
| Hussain, 2025 | DeepSeek | The standard classification accuracy | 54.5% | Average of three attending physicians: 51.5% | NA |
|  | ChatGPT‑01 |  | 63.6% |  |  |
|  | ChatGPT‑4.0 |  | 63.6% |  |  |
|  | Qwen 2.5 MAX |  | 54.5% |  |  |
| Delsoz, 2023 | ChatGPT 3.5 | The standard classification accuracy | 72.7% | Resident 1: 54.5% | NA |
|  |  |  |  | Resident 2: 72.7% |  |
|  |  |  |  | Resident 3: 72.7% |  |
