## Supplementary material for "Artificial Intelligence Powered Research Automation (AIPRA) Versus Human Expert: A Two-Arm Ophthalmology Comparative Study": AIRPA Manuscript

Manuscript 2

**Introduction**

Glaucoma, a leading cause of irreversible blindness globally, affects millions and poses significant challenges to healthcare systems due to its progressive and often asymptomatic nature [1]. Early diagnosis and intervention are critical to preventing vision loss, yet disparities in access to specialized care persist, particularly in low-resource settings [2]. Recent advancements in artificial intelligence (AI), particularly large language models (LLMs), offer promising tools to augment glaucoma diagnosis by analyzing multimodal data, including clinical notes, imaging, and patient histories [3]. These models have demonstrated potential in streamlining diagnostic workflows, improving accuracy, and bridging gaps in care delivery, though their integration into clinical practice remains an area of active research [4].

While LLMs show promise, their application in glaucoma diagnosis presents specific challenges and opportunities. For instance, studies reveal variability in diagnostic performance across models, with some achieving specialist-level accuracy in predicting glaucoma progression [5] while others struggle with interpretability and generalizability [6]. Key sub-problems include algorithmic bias, the need for domain-specific fine-tuning, and the integration of multimodal data such as optical coherence tomography (OCT) and visual field tests [7]. Controversies also persist regarding the reliability of LLMs in real-world settings, with some studies reporting high rates of diagnostic errors or "hallucinations" when interpreting fundus images [8]. Additionally, the development of specialized frameworks like GlaucoRAG, which leverages retrieval-augmented generation, highlights efforts to improve model precision by incorporating curated medical literature [4]. These advancements underscore the need for rigorous validation to ensure clinical utility [9].

The aim of this systematic review is to evaluate the current evidence on LLMs in glaucoma diagnosis, synthesizing their diagnostic accuracy, limitations, and potential for clinical integration. By addressing gaps in the literature—such as disparities in model performance and the lack of standardized evaluation metrics—this review seeks to inform future research and policymaking [2]. The findings will be particularly relevant for clinicians, researchers, and developers working toward equitable, AI-enhanced glaucoma care [10].

**Methods**

Search Strategy

A systematic search was conducted across multiple databases to identify relevant studies evaluating the application of large language models (LLMs) in glaucoma diagnosis. The search strategy included the following terms for PubMed:

("large language model*"[Title/Abstract] OR "LLM"[Title/Abstract] OR "LLMs"[Title/Abstract] OR "generative AI"[Title/Abstract] OR "GPT"[Title/Abstract] OR "ChatGPT"[Title/Abstract] OR "natural language processing"[MeSH Terms] OR "NLP"[Title/Abstract]) AND ("glaucoma"[MeSH Terms] OR "glaucoma"[Title/Abstract] OR "open-angle glaucoma"[Title/Abstract] OR "angle-closure glaucoma"[Title/Abstract] OR "ocular hypertension"[Title/Abstract] OR "glaucoma diagnosis"[Title/Abstract] OR "glaucoma management"[Title/Abstract])

Screening

Reviewers screened titles and abstracts for relevance. Full-text screening was then conducted to assess eligibility based on predefined inclusion and exclusion criteria.

Inclusion and Exclusion Criteria

Inclusion Criteria:

1. Studies evaluating LLMs (e.g., ChatGPT, GPT-4, BioBERT) in glaucoma diagnosis or management.

2. Studies reporting diagnostic accuracy, sensitivity, specificity, or other performance metrics.

3. Peer-reviewed original research articles.

Exclusion Criteria:

1. Studies not focused on glaucoma diagnosis or management.

2. Review articles, editorials, or conference abstracts without original data.

3. Studies lacking quantitative performance evaluation of LLMs.

Data Extraction

Data extraction was performed for the following fields from each included study: Primary Study Objective; Secondary Study Objectives; Study Design; Dataset Description; Dataset Size; LLM Model Used; Comparison Methods; Primary Outcome Measure; Secondary Outcome Measures; Sensitivity, Specificity, Accuracy, AUC (Area Under Curve); Statistical Significance. Limitations

The initial search resulted in 112 articles, where 83 articles were excluded after title and abstract screening. After full text screening, another 12 articles were excluded (supplementary material). A total of 18 articles met the inclusion criteria and were eventually included in the systematic review.

**Results**

**Overview of Included Studies**

This systematic review included 18 studies evaluating the application of large language models (LLMs) in glaucoma diagnosis. The studies varied in design, with retrospective cohort studies (n=5), evaluation/diagnostic accuracy studies (n=7), prospective observational studies (n=3), and cross-sectional studies (n=3). Sample sizes ranged from 11 to 3,170 eyes, with most studies utilizing ChatGPT variants (GPT-3.5, GPT-4, GPT-4o, GPT-4V) alongside other models such as BioBERT, RoBERTa, DistilBERT, DeepSeek, and Google Gemini. Data sources included electronic health records (EHRs), clinical case reports, visual field (VF) tests, fundus images, and standardized ophthalmology datasets (e.g., OHTS, REFUGE, ACRIMA).

**Diagnostic Accuracy of LLMs in Glaucoma Detection**

Several studies assessed LLMs' ability to diagnose glaucoma from text-based clinical notes, fundus images, and structured datasets.

- Text-Based Diagnosis:

- Hu & Wang (2022) evaluated BERT-based models on EHR notes, reporting AUROCs of 0.60–0.69, with BioBERT performing best (AUROC=0.69).

- Delsoz et al. (2023) found ChatGPT-3.5 correctly diagnosed 72.7% (8/11) of glaucoma cases, comparable to senior ophthalmology residents.

- Hussain et al. (2025) reported ChatGPT-4o achieved 84.9% accuracy in diagnosing glaucoma from case reports, outperforming Qwen 2.5 MAX (54.5%) and DeepSeek-V3 (54.5%).

- Image-Based Diagnosis:

- AlRyalat et al. (2024) tested ChatGPT-4 on 1,200 fundus images, achieving 50% sensitivity and 94.44% specificity, though with low reproducibility.

- Jalili et al. (2025) evaluated GPT-4V on 300 fundus images, showing sensitivity of 0.71–0.92 and specificity of 0.65–0.74 across datasets (ACRIMA, ORIGA, RIM-One).

- Gupta & Al-Kazwini (2024) found ChatGPT-4.0 correctly diagnosed only 33.3% (4/12) of retinal diseases, including glaucoma.

- Multimodal Inputs:

- Pan & Hastings (2025) combined saliency maps with clinical metadata, achieving 77.18% accuracy in ocular disease explanations.

- Mihalache et al. (2024) reported ChatGPT-4 had 61% accuracy on glaucoma-related image-based questions, lower than non-image questions (70% overall).

**Performance in Surgical Decision-Making**

Three studies evaluated LLMs' ability to recommend glaucoma surgeries. Carlà et al. (2025) tested ChatGPT-3.5 on 60 surgical cases, finding 78% agreement with specialists in "ordinary" cases but only 65% in challenging cases. Carlà et al. (2024) compared ChatGPT-4 and Google Gemini, reporting 58% vs. 32% agreement with specialists, respectively. Lastly, Tan (2025) assessed GPT-4o on VF test reports, showing 96.7% accuracy in classifying glaucoma but only 73.3% accuracy in defect-type identification.

**Agreement with Human Experts**

Most studies evaluating large language models (LLMs) in ophthalmology compared their outputs with ophthalmologists or standardized diagnostic references. Zhang et al. (2024) reported that GPT-4o achieved 55% diagnostic accuracy, significantly lower than human ophthalmologists (p < 0.05). In contrast, GlaucoRAG (Aminan et al., 2025) outperformed GPT-4.5-PREVIEW, achieving 91.2% accuracy compared with 84.4% on glaucoma-specific questions. Similarly, Ming et al. (2024) found that GPT-4.0 matched the performance of ophthalmology residents in 50% of glaucoma triage cases, whereas GPT-3.5 lagged behind with only 20% accuracy.

**Predictive Modeling for Glaucoma Progression**

Two studies used LLMs to predict glaucoma progression from clinical data. Hu & Wang (2022) reported BioBERT achieved the highest AUROC (0.69) in predicting surgical progression. Huang et al. (2024) tested ChatGPT-4.0 on OHTS data, yielding an AUC of 0.67, while ChatGPT-3.5 performed slightly worse (AUC = 0.62).

Predictive performance varied by model architecture, with domain-specific fine-tuning (e.g., BioBERT) improving accuracy.

**Limitations and Variability**

Common limitations included small sample sizes (e.g., Gupta & Al-Kazwini (2024): n=12); lack of multimodal integration (e.g., Carlà et al. (2025) noted ChatGPT’s inability to interpret fundus images); reproducibility issues, such as AlRyalat et al. (2024) reporting inconsistent responses to the same fundus images; and dataset bias, with most studies using Western or Chinese populations (e.g., REFUGE, OHTS).

**Summary of Quantitative Findings**

| **Outcome** | **Model** | **Performance** | **Study** |
| --- | --- | --- | --- |
| **Text-Based Diagnosis** | BioBERT | AUROC=0.69 | Hu & Wang (2022) |
| **Image-Based Diagnosis** | GPT-4V | Sensitivity=0.71–0.92 | Jalili et al. (2025) |
| **Surgical Decision-Making** | ChatGPT-3.5 | 78% agreement (ordinary cases) | Carlà et al. (2025) |
| **Agreement with Specialists** | GlaucoRAG | 91.2% accuracy (BCSC questions) | Aminan et al. (2025) |
| **Multimodal Interpretation** | GPT-4o | 96.7% VF defect classification | Tan (2025) |

**Discussion**

The systematic review included 18 studies evaluating the role of large language models (LLMs) in glaucoma diagnosis, progression prediction, and clinical decision support. Key findings revealed that LLMs demonstrated variable performance across different tasks. In regard to diagnostic Accuracy, several studies reported moderate to high accuracy in diagnosing glaucoma from clinical notes, visual field reports, and fundus images. For instance, models like BioBERT and GPT-4 achieved AUROCs between 0.60–0.76 for predicting glaucoma progression from electronic health records (EHRs), while multimodal LLMs showed 73–96.7% accuracy in interpreting visual field defects. However, performance varied significantly based on input data type, with image-based interpretations generally less reliable than text-based analyses.

In terms of clinical Decision Support LLMs like ChatGPT-3.5 and ChatGPT-4 showed promise in suggesting surgical interventions, with 65–83% concordance with specialists in routine cases but lower accuracy in complex scenarios. Retrieval-augmented models (e.g., GlaucoRAG) outperformed general-purpose LLMs in glaucoma-specific question-answering (91.2% accuracy). For prognostic Applications, some studies explored LLMs for predicting glaucoma conversion from ocular hypertension, achieving AUCs of 0.62–0.76, though performance lagged behind traditional machine learning models in some cases.

The findings align with broader trends in AI applications for glaucoma, though LLMs introduce unique opportunities and challenges. Glaucoma remains a leading cause of irreversible blindness globally [1], yet early detection is often hindered by asymptomatic progression. While LLMs show potential in automating risk stratification, their diagnostic accuracy (e.g., 50–96.7% in this review) is still below the near-perfect performance of specialized deep learning models for optical coherence tomography (OCT) analysis [2]. This suggests LLMs may complement, but not yet replace, imaging-based AI tools. Prior reviews highlight the superiority of convolutional neural networks (CNNs) for OCT-based glaucoma detection [2], whereas LLMs excel in synthesizing textual data (e.g., clinical notes, research literature). The current review supports this dichotomy, as LLMs struggled with image interpretation unless augmented with retrieval systems (e.g., GlaucoRAG) or structured data pipelines. Unlike traditional machine learning, which focuses on specific tasks like visual field analysis [3], LLMs offer versatility in generating explanations and summaries. However, their inconsistency in complex cases mirrors broader concerns about AI reliability in ophthalmology [2].

There are several limitations of this review, including heterogeneity of Studies: Varied evaluation metrics (e.g., accuracy, AUROC, F1 scores) and datasets (EHRs, fundus images, synthetic cases) limited direct comparisons. Another limitation is the focus on early-stage models, as most studies evaluated prototype LLMs (e.g., ChatGPT-3.5/4) rather than clinically validated systems. Performance may improve with fine-tuned, domain-specific models. Lastly, studies predominantly used retrospective data from single centers or curated datasets (e.g., REFUGE), which may not reflect real-world diversity in patient populations or imaging quality.

The implications of using large language models (LLMs) in ophthalmology extend across clinical, research, and regulatory domains. In clinical practice, LLMs hold promise for tasks such as triaging high-risk patients through electronic health record (EHR) analysis or summarizing diagnostic reports; however, consistent human oversight is essential given their variable performance. Research directions should focus on multimodal fusion, particularly integrating LLMs with convolutional neural networks (CNNs) for joint image–text analysis as highlighted in AI-OCT studies, as well as on mitigating bias by addressing dataset limitations, such as the underrepresentation of diverse populations, potentially through federated learning approaches. Furthermore, real-time validation in prospective studies comparing LLMs to clinicians in dynamic settings is needed to assess reliability. Finally, the development of ethical and regulatory frameworks remains critical to ensure accountability and transparency in LLM-generated diagnoses, as underscored in recent AI reviews.
